# Assessment of Bacteriological Quality and Associated Risk Factors of Raw Milk Marketed in Nyagatare district, Rwanda

**DOI:** 10.1101/2023.07.24.23293027

**Authors:** Niyigena Emmanuel, Kwizera Enock

**Author notes:** Corresponding author’s.

## Abstract

Milk has tremendous nutritional value for both people and animals, but because it also contains water, many different kinds of microbes can develop in it. Dairy products are still sold in both regular and unofficial marketplaces in Rwanda. Significant dangers to human health are associated with milk sold in unregulated markets. The aim of the present study was to assess the bacteriological quality and associated risk factors of raw milk marketed in Nyagatare district, Rwanda. 126 samples were collected among milk transporters from 14 sectors of Nyagatare district, Rwanda, where 9 samples were taken from each sector and analyzed in veterinary microbiology lab of University of Rwanda, Nyagatare campus. The bacterial analysis was based on carrying out total bacterial count, Coliform count, enumeration of Staphylococcus species and enumeration of Salmonella species. Milk transportes were also asked about risk factors associated with bacterial contamination of raw milk. The total bacterial count, Coliform count and enumerated Staphylococcus species had range from 7.8 × 10^4^ -80 × 10^4^ cfu/mL, 24 × 10^1^ and 80 × 10^1^ cfu/mL and 20.8 × 10^1^ and 211 × 10^1^ cfu/mL respectively and highest prevalence of Salmonella to be 1.58 %. The level of significant was not significant between various bacterial level and several risk factors except for total bacterial count and knowledge about diseases transmitted by raw milk at p < 0.05. It is, therefore, highly recommended that all concerned parties should increase awareness about diseases transmitted by raw milk among milk transporters in Nyagatare district among others.

## 1 Introduction

Milk and dairy products have been a reliable source of nutrition and comfort for generations, contributing significantly to global nutrition and economies. However, milk can also pose health risks due to various contaminants, making food safety a critical concern [1]. In Rwanda, the dairy industry plays a vital role in the country’s sustainable development objectives, offering economic opportunities and improved food security. Nyagatare district, one of leading milk producer in Rwanda, faces challenges in maintaining the quality and safety of milk due to traditional handling practices and inadequate knowledge among small-holder farmers [2]. This study aimed at assessing the bacteriological quality of raw milk marketed in Nyagatare district and identifying the associated risk factors. The research objectives include evaluating the level of bacterial contamination and determining the factors influencing such contamination. The study aims to answer the research questions concerning the bacteriological contamination level and the practices of milk transporters contributing to contamination. Milk provides essential nutrients but also creates favourable conditions for microbial growth [3]. Therefore, assessing the bacteriological quality and understanding the influencing factors are crucial for establishing effective control measures. The study was conducted over a four-month period, collecting milk samples from various sources and locations within Nyagatare district, providing a diverse sample set.

In terms of milk production, Rwanda has three main dairy production systems: zero grazing, open grazing, and semi-grazing [4]. The volume of milk produced in the country has significantly increased over the years, reaching 999,976 MT in 2021. The national cattle herd consists of 1.5 million cows, with more than half being genetically improved dairy breeds. Hybrid cows resulting from artificial insemination services have become the dominant breed. However, the average milk yields are still below the genetic potential of the cows [5]. Regarding milk collection, smallholder farmers in Rwanda transport milk to Milk Collection Centers or market outlets using various means such as walking, bicycles, motorcycles, and trucks [4]. Throughout the fiscal year 2021/22, a grand total of 85,223,665 liters of milk were directed through the 132 operational Milk collection centers. The collected milk is then supplied to processing plants. Milk Collection Centers operate throughout the day or twice a day to accommodate different milking times [5]. The milk market in Rwanda consists of both informal and formal sectors. Approximately 85% of the milk is handled by informal traders, where quality and food safety cannot be guaranteed. These informal traders include milk collectors, hawkers, and traders who sell raw milk through designated sales points such as milk kiosks. On the other hand, 15% of the milk is handled by formal traders who adhere to quality and food safety standards [6]. The formal milk market is regulated by a Ministerial order that requires traders to obtain licenses and sell tested and certified milk. Supermarkets, milk shops, and milk zones are the main sales points in the formal milk market, offering different types of milk products [4].

In terms of food safety, raw milk can be contaminated with various milk-borne pathogens, including coliforms, Staphylococcus species, and Salmonella. Coliform bacteria, which can indicate the presence of enteric pathogens, are commonly detected using bacteriological media containing lactose. Staphylococcus species, including Staphylococcus aureus, can cause foodborne illnesses and spoilage in dairy products [7]. Detection and identification of Staphylococcus species can be done using microscopy, biochemical tests, and molecular techniques. Salmonella, a widespread foodborne pathogen, requires culture-based detection and isolation procedures using selective media. Ensuring food safety in relation to raw milk involves addressing factors such as animal health, farm management practices, environmental hygiene, and temperature control [3].

## 2 Research Methodology

This study was conducted in the Nyagatare district of Rwanda. This district is one of leading milk producer in the country and is known for its agricultural practices, with a focus on cattle keeping. It is an expansive region with grassy plains and low hills [2, 8]. The study design employed a cross-sectional approach. Bacteriological analysis was performed on milk samples collected from milk transporters in Nyagatare district to determine the bacterial level of raw milk marketed in the area. Additionally, a survey questionnaire was used to gather data from milk transporters on factors influencing the level of bacterial contamination in raw milk. The target population for the study consisted of milk transporters to the Milk Collection Centers (MCC) in Nyagatare district. The sample size of 126 milk transporters was determined using an online sample size calculator. Data collection comprised two parts: a questionnaire-based survey and bacteriological quality analysis.

The structured questionnaire was used to collect information from milk transporters, focusing on factors influencing the level of bacterial contamination in raw milk. The questionnaire included pre-coded response choices and was administered using the Kobo toolbox, an online survey data collection tool. Raw milk samples were collected from milk transporters in Nyagatare district. Aseptic techniques were used to collect 30-milliliter milk samples directly from participant containers, which were then placed into sterile screw cups. The samples were labelled, stored in a 4°C icebox, and transported to the microbiology laboratory for analysis [9]. Bacteriological analysis involved several steps. Total bacterial count and Coliform count were determined using plate count agar and violet red bile agar, respectively [10]. Staphylococcus species were enumerated using Baird Parker Agar [11], while Salmonella species were detected through pre-enrichment and selective enrichment techniques, followed by incubation and plate streaking on xylose lysine deoxycolate (XLD) agar [6].

Data analysis was conducted using SPSS version 27. Descriptive statistics, such as frequencies and percentages, were used to analyze quantitative data from the survey questionnaires and microbial analysis. The relationship between bacterial levels and risk factors was assessed using the Multinomial Logistic Regression test with Pearson’s rank correlation coefficient. The bacteriological analysis results were also compared to the standards set by the Rwanda Standard Board for raw milk. Quality control measures were implemented throughout the study to ensure the validity of the obtained data. These included pretesting the questionnaire, using calibrated equipment, checking the quality and shelf life of reagents and materials, and following the manufacturer’s instructions for media preparation and sterilization.

## 3 Results and Discussion

Results of the total bacterial count (n = 126) analyses revealed the total bacterial count mean among milk transporters in Nyagatare district to vary between 7.8 × 10^4^ and 80 × 10^4^ cfu/mL as illustrated in **Table 1** and the majority of the milk samples met total bacterial count limit recommended by Rwanda standard board, where it recommends 2 × 10^6^ cfu/mL for raw milk and this is in range with East African Community standard, who recommends the same limit for raw milk, 2 × 106 cfu/mL [12]. The findings of the study are lower compared to the study of [13] who reported the mean TBC range between 5.3 × 10^5^ and 2.4 × 10^8^ cfu/mL for MCC in Rwanda. In addition, the findings of the study are lower compared to the study of [6] who reported the mean TBC range between 2.3 × 10^5^ cfu/ml to 9.1 × 10^5^ cfu/ml among raw milk at kiosks in Ines Ruhengeri. The high TBC in milk from MCC and kiosks suggests proliferation or recontamination of milk by bacteria during transportation.

**Table 1:** Total bacterial count among milk transporters.

| Location | Mean ($\times 10^4$ cfu/mL) | SD | Min ($\times 10^4$ cfu/mL) | Max ($\times 10^4$ cfu/mL) |
| --- | --- | --- | --- | --- |
| Nyagatare sector | 32 | 642,564 | 4.7 | 203 |
| Rwempasha sector | 14.9 | 74,689 | 6.1 | 29.1 |
| Tabagwe sector | 7.8 | 56,289 | 2.6 | 20.2 |
| Rwimiyaga sector | 58 | 965,263 | 2.9 | 243.6 |
| Matimba sector | 11.5 | 75,023 | 3.4 | 24.1 |
| Musheli sector | 9.3 | 70,251 | 2.8 | 24.1 |
| Rukomo sector | 12.6 | 57,052 | 6.6 | 21.6 |
| Karama sector | 8.9 | 62,862 | 3 | 22.2 |
| Gatunda sector | 36 | 694,292 | 3.1 | 220 |
| Karangazi sector | 11.7 | 70,326 | 3.3 | 24 |
| Katabagemu sector | 10.8 | 58,180 | 2.7 | 20.4 |
| Kiyombe sector | 80 | 1,101,584 | 3.2 | 237 |
| Mukama sector | 52 | 916,827 | 2.9 | 215 |
| **Mimuli sector** | 11.4 | 53,814 | 5.2 | 21.5 |

Results of the Coliform count analyses revealed the Coliform count mean among milk transporters in Nyagatare district to vary between 24 × 10^1^ and 80 × 10^1^ cfu/mL as illustrated in **Table 2**. The majority of the milk samples met Coliform count limit recommended by Rwanda standard board, where it recommends 5 × 10^4^ cfu/mL for raw milk and this is in line with recommended standard by East African Community where it also recommends 5 × 10^4^ cfu/mL for raw milk [12]. This study findings are higher compared to the study by [6] who reported 1.8 × 10^2^ to 2.9 × 10^2^ cfu/ml among raw milk in kiosks in Ines Ruhengeri.

**Table 2:** *Coliform* count among milk transporters.

| Location | Mean ($\times 10^1$ cfu/mL) | SD | Min ($\times 10^1$ cfu/mL) | Max ($\times 10^1$ cfu/mL) |
| --- | --- | --- | --- | --- |
| **Nyagatare sector** | 80 | 800 | 80 | 80 |
| **Rwempasha sector** | 0 | 0 | 0 | 0 |
| **Tabagwe sector** | 78 | 780 | 78 | 78 |
| **Rwimiyaga sector** | 24 | 42.426 | 21 | 27 |
| **Matimba sector** | 31 | - | 3 | 31 |
| **Musheli sector** | 0 | 0 | 0 | 0 |
| **Rukomo sector** | 66 | - | 66 | 66 |
| **Karama sector** | 80 | - | 80 | 80 |
| **Gatunda sector** | 0 | 0 | 0 | 0 |
| **Karangazi sector** | 61 | - | 61 | 61 |
| **Katabagemu sector** | 0 | 0 | 0 | 0 |
| **Kiyombe sector** | 0 | 0 | 0 | 0 |
| **Mukama sector** | 0 | 0 | 0 | 0 |
| **Mimuli sector** | 0 | 0 | 0 | 0 |

Results of the enumerated Staphylococcus species analyses revealed the enumerated Staphylococcus species mean among milk transporters in Nyagatare district to vary between 20.8 × 10^1^ and 211 × 10^1^ cfu/mL as illustrated in **Table 3**. The majority of the milk samples met Staphylococcus species count limit recommended by RSB, where it recommends 2 × 10^3^ cfu/mL for raw milk and this is in line with recommended standard by East African Community where it also recommends 2 × 10^3^ cfu/mL for raw milk [12]. This is in range with study by (Alice et al., 2021) who reported 1.8 × 10^2^ to 2.2 × 10^2^ cfu/ml among milk samples in kiosks in Ines Ruhengeri. Finding Staphylococcus species pose a health safety problem to milk consumer in Nyagatare district as the study by [4] reported that milk in Rwanda, is consumed as raw among other ways.

**Table 3:** Enumerated Staphylococcus species among milk transporters.

| Location | Mean ($\times 10^1$ cfu/mL) | SD | Min ($\times 10^1$ cfu/mL) | Max ($\times 10^1$ cfu/mL) |
| --- | --- | --- | --- | --- |
| **Nyagatare sector** | 0 | 800 | 0 | 0 |
| **Rwempasha sector** | 0 | 0 | 0 | 0 |
| Tabagwe sector | 23.4 | 780 | 23.4 | 23.4 |
| Rwimiyaga sector | 67 | 42.426 | 11 | 123 |
| Matimba sector | 211 | - | 211 | 211 |
| Musheli sector | 0 | 0 | 0 | 0 |
| Rukomo sector | 106 | - | 43 | 170 |
| Karama sector | 0 | - | 0 | 0 |
| Gatunda sector | 0 | 0 | 0 | 0 |
| Karangazi sector | 61 | - | 36 | 78 |
| Katabagemu sector | 0 | 0 | 0 | 0 |
| Kiyombe sector | 20.8 | 0 | 20.8 | 20.8 |
| Mukama sector | 0 | 0 | 0 | 0 |
| Mimuli sector | 0 | 0 | 0 | 0 |

Results of the enumerated Salmonella analyses revealed that the highest prevalence of enumerated Salmonella among milk transporters in Nyagatare district was in Rwempasha and Matimba sector and it was estimated to be 1.58 % each as illustrated in **Table 4**. The majority of the milk samples met salmonella species count limit recommended by Rwanda standard board, where it recommends absent for raw milk and this is in line with recommended standard by East African Community where it also recommends absent for raw milk [12]. The prevalence of salmonella species found in this study is lower compared to the study of [14] who reported the prevalence of 5.2% along Rwandan milk and dairy chain.

**Table 4:** Enumerated Salmonella species among milk transporters.

| Variable | Category | Frequency (n=126) | Percentage (%) |
| --- | --- | --- | --- |
| Nyagatare sector | *Salmonella species* found | 0 | 0 |
|  | No *Salmonella species* found | 9 | 7.142 |
| Rwempasha sector | *Salmonella species* found | 2 | 1.58 |
|  | No *Salmonella species* found | 7 | 5.55 |
| Tabagwe sector | *Salmonella species* found | 1 | 0.79 |
|  | No *Salmonella species* found | 8 | 6.35 |
| Rwimiyaga sector | *Salmonella species* found | 0 | 0 |
|  | No *Salmonella species* found | 9 | 7.142 |
| Matimba sector | *Salmonella species* found | 2 | 1.58 |
|  | No *Salmonella species* found | 7 | 5.55 |
| Musheli sector | *Salmonella species* found | 0 | 0 |
|  | No *Salmonella species* found | 9 | 7.142 |
| Rukomo sector | *Salmonella species* found | 1 | 0.79 |
|  | No *Salmonella species* found | 8 | 6.35 |
| Karama sector | *Salmonella species* found | 0 | 0 |
|  | No *Salmonella species* found | 9 | 7.142 |
| Gatunda sector | *Salmonella species* found | 0 | 0 |
|  | No *Salmonella species* found | 9 | 7.142 |
| Karangazi sector | *Salmonella species* found | 1 | 0.79 |
|  | No *Salmonella species* found | 8 | 6.35 |
| Katabagemu sector | *Salmonella species* found | 0 | 0 |
|  | No *Salmonella species* found | 9 | 7.142 |
| Kiyombe sector | *Salmonella species* found | 0 | 0 |
|  | No *Salmonella species* found | 9 | 7.142 |
| **Mukama sector** | *Salmonella species* found | 0 | 0 |
|  | No *Salmonella species* found | 9 | 7.142 |
| **Mimuli sector** | *Salmonella species* found | 0 | 0 |
|  | No *Salmonella species* found | 9 | 7.142 |

Results of the survey revealed that, of all 126 participants, 16 respondents (12.7%) said that after milking, they transport milk to final destination for more than 2 hours after milking, and they do not keep it refrigerated as illustrated **Table 5**. This agree with the study of [15] who reported that after milking in north-western part of Rwanda, milk is kept at farm for more than 5 hours before taken by milk transporters among others or even when taken after milking, it can be taken to final destination for more than 2 hours and it is not kept in refrigeration condition. This may be led proliferation of bacterial to unsafe level for consumption as reported in the study by [16]. Again, knowledge about diseases transmitted by raw milk was the only factor that was significantly (P < 0.05) associated with the level of total bacterial count among milk transporters in Nyagatare district as illustrated **Table 6**. This agree with the study of [17] who reported that the awareness of the diseases that can be transmitted by raw milk, is essential in reducing the hazards associated with drinking raw milk.

**Table 5:**
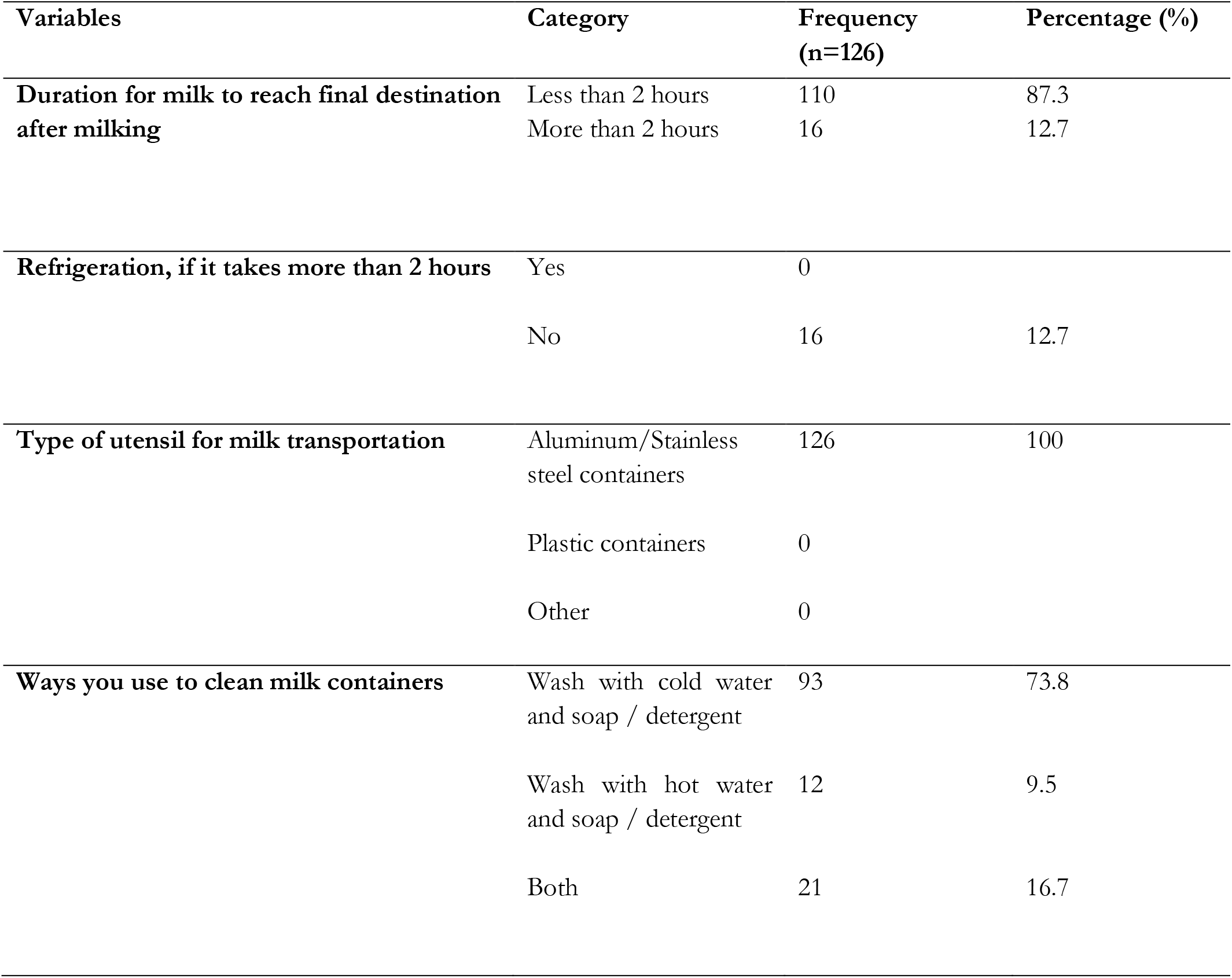
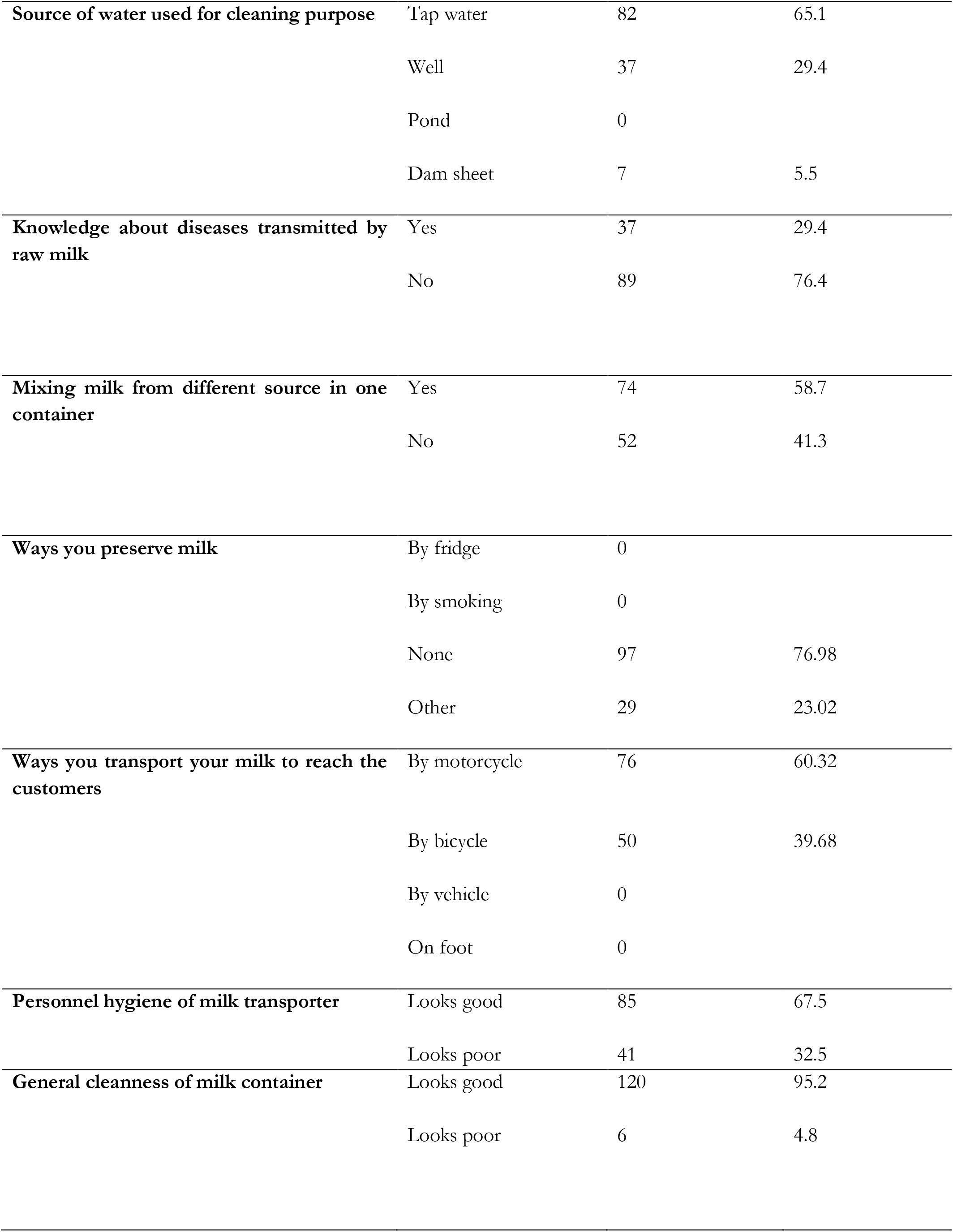
The factors influencing the level of bacterial contamination.

| Variables | Category | Frequency<br>(n=126) | Percentage (%) |
| --- | --- | --- | --- |
| **Duration for milk to reach final destination after milking** | Less than 2 hours | 110 | 87.3 |
|  | More than 2 hours | 16 | 12.7 |
| **Refrigeration, if it takes more than 2 hours** | Yes | 0 |  |
|  | No | 16 | 12.7 |
| **Type of utensil for milk transportation** | Aluminum/Stainless steel containers | 126 | 100 |
|  | Plastic containers | 0 |  |
|  | Other | 0 |  |
| **Ways you use to clean milk containers** | Wash with cold water and soap / detergent | 93 | 73.8 |
|  | Wash with hot water and soap / detergent | 12 | 9.5 |
|  | Both | 21 | 16.7 |
| **Source of water used for cleaning purpose** | Tap water | 82 | 65.1 |
|  | Well | 37 | 29.4 |
|  | Pond | 0 |  |
|  | Dam sheet | 7 | 5.5 |
| **Knowledge about diseases transmitted by raw milk** | Yes | 37 | 29.4 |
|  | No | 89 | 76.4 |
| **Mixing milk from different source in one container** | Yes | 74 | 58.7 |
|  | No | 52 | 41.3 |
| **Ways you preserve milk** | By fridge | 0 |  |
|  | By smoking | 0 |  |
|  | None | 97 | 76.98 |
|  | Other | 29 | 23.02 |
| **Ways you transport your milk to reach the customers** | By motorcycle | 76 | 60.32 |
|  | By bicycle | 50 | 39.68 |
|  | By vehicle | 0 |  |
|  | On foot | 0 |  |
| **Personnel hygiene of milk transporter** | Looks good | 85 | 67.5 |
|  | Looks poor | 41 | 32.5 |
| **General cleanness of milk container** | Looks good | 120 | 95.2 |
|  | Looks poor | 6 | 4.8 |

**Table 6:**
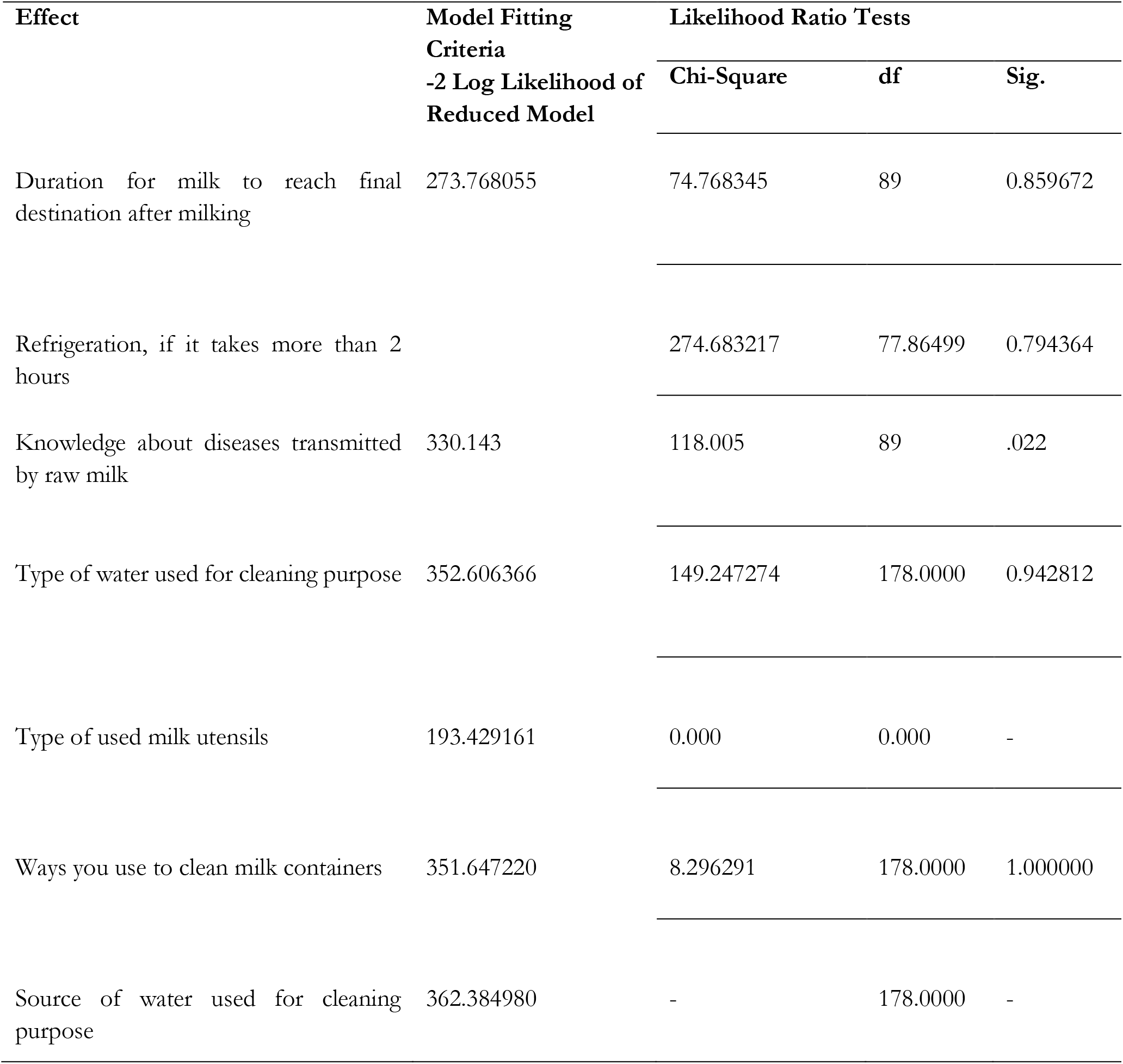
Multinomial Logistic Regression of bacterial level and associated risk factors.

## 4 Conclusions

This study provided information on the level of the bacterial quality and associated influencing risk factors of raw milk marketed in Nyagatare district among milk transporters. The study documented a relatively good quality raw milk and few associated influencing risk factors in regard to bacterial levels. The bacterial level found are associated with various factors particularly knowledge about diseases transmitted through raw milk. As a result, it is critical to raise the awareness about diseases transmitted through raw milk among milk transporters in Nyagatare distict. Moreover, because of the discovered milk contamination particularly, *Coliforms, Staphylococcus species* and *Salmonella*, it was determined that the milk among milk transporters posed a public health risk and that it should not be consumed raw. To reduce such contamination and bacteriological load in raw milk, standard operating methods should be practiced, such methods include hand washing before handling milk cans and proper washing of milk cans with hot water and soaps. Furthermore, further studies are needed to isolate and characterize bacterial loads in raw milk marketed among milk transporters in Nyagatare district, Rwanda. Based on the findings of this study, several recommendations can be made. Firstly, there should be regular awareness campaigns aimed at educating milk transporters about the diseases that can be transmitted through raw milk and the preventive measures they can take. Secondly, the effectiveness of raw milk handling hygiene trainings among milk transporters should be improved by increasing their frequency and scope. Thirdly, it is crucial to emphasize the importance of personal hygiene among milk transporters and encourage them to maintain high standards in this regard. Lastly, public health authorities should play an active role by intervening in training initiatives and monitoring the implementation of good hygienic practices (GHP) throughout the entire milk value chain. By implementing these recommendations, we can enhance the safety and quality of raw milk and minimize the risk of disease transmission through consumption of raw milk in Nyagatare district.

## Data Availability

All data produced in the present work are contained in the manuscript

## 5 Declarations

### 5.1 Study Limitations

The study did not perform drug sensitivity testing of isolated milk pathogenic bacteria. The study was cross-sectional so missed out on understanding the effects of seasonality on milk microbial quality. Isolated bacteria were not typed to strain level using molecular methods. This study had no reference standards of bacterial strains to use in the lab like ATCC references.

### 5.2 Funding source

Funding for this project was provided by the University of Minnesota-University of Rwanda Global Animal Health Fellowship Program to support research collaboration between both institutions.

### 5.3 Competing Interests

I, the author, declare that there is no conflict of interest in this work.

## Notes

### Competing Interest Statement

The authors have declared no competing interest.

### Funding Statement

This study was funded by University of Minnesota - University of Rwanda Global Animal Health Fellowship Program

## References

[1] G. Kansal, A. Kumar, and V. Yadav, “Quality milk production for profitable dairying,” Vigyan Varta, vol. 3’ no. 1’ pp. 30–34, 2022.

[2] S. N. Garcia, J. P. M. Mpatswenumugabo, P. Ntampaka, S. Nandi, and J. S. Cullor, “A one health framework to advance food safety and security: An on-farm case study in the Rwandan dairy sector,” One Health, vol. 16, p. 100531, 2023.

[3] K. K. Dash et al., “A comprehensive review on heat treatments and related impact on the quality and microbial safety of milk and milk-based products,” Food Chemistry Advances, vol. 1, p. 100041, 2022.

[4] N. Habiyaremye, E. A. Ouma, N. Mtimet, and G. A. Obare, “A Review of the Evolution of Dairy Policies and Regulations in Rwanda and Its Implications on Inputs and Services Delivery,” (in English), Frontiers in Veterinary Science, Policy and Practice Reviews vol. 8, 2021-July-23 2021, doi: 10.3389/fvets.2021.611298.

[5] MINAGRI, “MINAGRI ANNUAL REPORT 2021-2022,” Rwanda Ministry of Agriculture and Animal Resources (MINAGRI), Rwanda, 2022. [Online]. Available: https://www.minagri.gov.rw/index.php?eID=dumpFile&t=f&f=56427&token=53172b32acc651a19a341690971cb577612cbf8f

[6] I. I. Alice, N. Joseph, H. Innocent, and D. A. Celise, “Assessment of microbiological quality of raw milk produced and commercialized around INES Ruhengeri, Musanze district, Rwanda,” African Journal of Biological Sciences, vol. 3, no. 4, 2021.

[7] S. Z. Abbas et al., “Microbiological assessment of raw milk available in the metropolitan city of Sindh, Karachi-Pakistan: Microbiological Assessment of Raw Milk,” Pakistan Journal of Health Sciences, pp. 220–224, 2022.

[8] E. Mazimpaka, F. Mbuza, T. Michael, E. N. Gatari, E. Bukenya, and O.-A. James, “Current status of cattle production system in Nyagatare District-Rwanda,” Tropical animal health and production, vol. 49, pp. 1645–1656, 2017.

[9] A. Aliyo and Z. Teklemariam, “Assessment of Milk Contamination, Associated Risk Factors, and Drug Sensitivity Patterns among Isolated Bacteria from Raw Milk of Borena Zone, Ethiopia,” Journal of Tropical Medicine, vol. 2022, p. 3577715, 2022/06/20 2022, doi: 10.1155/2022/3577715.

[10] N. C. Sharma, B. Bais, L. Tak, J. Singh, and Y. Singh, “Microbial quality evaluation of aloe vera and coconut water based whey beverages prepared from camel and goat milk,” Pharma Innov, vol. 10, pp. 137–140, 2021.

[11] R. H. Perez and A. E. Ancuelo, “Isolation and Characterization of Staphylococcus spp. and the Unintended Discovery of Non–Staphylococcal Strains Associated with Bovine Mastitis in Region IV-A, Philippines,” Philippine Journal of Science, vol. 151, no. 5, 2022.

[12] EAC. “EAST AFRICAN STANDARD: Raw cow milk — Specification.” East African Community. https://law.resource.org/pub/eac/ibr/eas.67.2006.html (accessed 1 December, 2022).

[13] J. B. Ndahetuye et al., “MILK Symposium review: Microbiological quality and safety of milk from farm to milk collection centers in Rwanda,” Journal of Dairy Science, vol. 103, no. 11, pp. 9730–9739, 2020, doi: 10.3168/jds.2020-18302.

[14] O. Kamana, S. Ceuppens, L. Jacxsens, A. Kimonyo, and M. Uyttendaele, “Microbiological quality and safety assessment of the Rwandan milk and dairy chain,” (in eng), no. 1944-9097 (Electronic), 2014.

[15] J. Mpatswenumugabo, L. Bebora, G. Gitao, V. Mobegi, B. Iraguha, and B. Shumbusho, “Assessment of bacterial contamination and milk handling practices along the raw milk market chain in the north-western region of Rwanda,” 11/30 2019.

[16] M. Doyle, S. Garcia, E. Bahati, D. Karamuzi, J. Cullor, and S. Nandi, “Microbiological analysis of raw milk in Rwanda,” African Journal of Food Science and Technology, vol. 6, pp. 141–143, 09/01 2015, doi: 10.14303/ajfst.2015.053.

[17] R. Fagnani, L. A. Nero, and C. P. Rosolem, “Why knowledge is the best way to reduce the risks associated with raw milk and raw milk products,” (in eng), no. 1469-7629 (Electronic), 2021.

